# Using Electronic Health Records to Understand Health in Emerging Communities

**DOI:** 10.1101/2025.06.27.25330411

**Authors:** Sarah Yeoh, John Licorish, Mai Stafford

## Abstract

**Background:** Ethnic inequalities in health and care are well-established. However, the lack of fine-grained data and small group sizes mean that the health and care needs of some ethnic groups are not well-quantified.

**Methods:** The North West London (NWL) Whole Systems Integrated Care (WSIC) database contains linked, deidentified records from General Practices (GPs), hospitals, and social care in NWL. Using this data, we examined health outcomes and healthcare utilisation of the Romanian and Somali communities in Brent.

**Results:** After age- and sex-standardisation, Somalis had higher prevalence of long-term conditions (LTCs), while Romanians had lower prevalence. Even after adjusting for prevalence of LTCs, the odds of having had an emergency department attendance, or hospital admission was higher for Somali adults, while Romanian adults had higher odds of having hospital admissions or avoidable emergency department attendances.

**Conclusion:** This work is the first to quantify health outcomes and utilisation of these communities in the UK, and demonstrate that routine healthcare data can be used to understand and quantify ethnic inequalities in health and care.

## 1 Introduction

Ethnic differences in health and care within the UK have been the subject of some interest for many decades [1–4]. However, such disparities were largely sidelined in policy work until the recent pandemic [5], which spotlighted the fact that ethnic minorities were disproportionately affected by Covid-19 in public media [6, 7], academic work [8–10], and government reports [11, 12]. This catalysed action to formally evaluate and address racial and ethnic inequality in the pandemic [13], and beyond the pandemic, to tackle the structural roots of these disparities.

Since then, the Core20PLUS5 framework by NHS England [14] represents an acknowledgment that health inequalities run deep through society, and a commitment to addressing the contribution of healthcare to these inequalities. Within the framework, the ‘PLUS’ population reflects a nuanced approach to addressing these inequities, by recognising that membership in marginalised groups can differ across the country, and effectively acknowledging that local authorities are best-placed to identify such groups, which can include ethnic minorities.

Brent is London and the UK’s second most ethnically diverse borough. Based on the latest census, 65% of the population identifies as being from a Black, Asian, or other minoritised ethnic group, and 56% of its residents were born overseas - the highest proportion in England and Wales. In particular, the Romanian and Somali communities within Brent are amongst the largest in the UK. Romanians comprise the second largest foreign-born community in Brent, and also its fastest-growing population since the 2011 Census [15]. Understanding the needs of these communities is therefore an imperative in addressing health inequalities within the borough.

However, quantitative research regarding the health and care outcomes and experiences of these two specific groups within the UK is scarce. Work on migrant Romanians is predominantly – if not exclusively – qualitative and sometimes groups them with Poles and Slovaks under the umbrella of ‘Eastern Europeans’ (e.g. [16, 17]), or more widely with the European Union A8 and A2 countries under ‘Central and Eastern European’ countries (e.g. [18, 19]). Within these wider groups, qualitative research has focused on healthcare instead of outcomes, and has highlighted language barriers to understanding information [17, 18, 20, 21] and encounters with healthcare staff, as well as experiences of discrimination [16]. While General Practice (GP) registration tends to increase with length of UK residence [19], transnational healthcare usage can be frequent [16–18].

Comparatively, there is more literature on the Somali community likely because Somali communities were established in the UK relatively earlier – products of the colonial era and Somali civil war [22]. Most of it, however, is qualitative, focusing on health beliefs and healthcare experiences. Research highlights poor awareness around health, in areas including diet [23], physical activity [24], healthy child body weight [25, 26], and available services such as smoking cessation services [27]. An area of especially poor health awareness is the measles, mumps and rubella (MMR) vaccine, which Somali communities tend to link to autism [28, 29], with poorer uptake as a consequence. Within the London borough of Tower Hamlets, 56% of Somali children had received the MMR1 vaccine by two years of age - compared to 94% across the borough [30]. Comparatively, reported uptake for diphteria-containing vaccines was higher - at around 85% at one year of age [31, 32], and around 88% at two years of age [32]. Notably, however, both rates were still lower than that for children of White British ethnicity [31, 32]. Similar to Romanians, Somalis reported feeling discriminated due to language but additionally also because of their skin colour [33]. Likely due to high numbers of Somali asylum seekers from the civil war, mental health is the subject of multiple studies. Specifically, mental health is poorly conceptualised by Somalis [34], with service usage being low [35] due to stigma [36–38].

In summary, there is a lack of quantitative evidence on the health status and healthcare activity in both Romanian and Somali communities. National survey samples tend to contain too few of either ethnicity, while routine health data has so far been under-utilised for research to describe health in these communities.

An obstacle to quantifying and addressing ethnic inequalities is the poor recording of ethnic data. Within the NHS, the recording of ethnic identity for ethnic minorities has often been inaccurate [39] and not always consistently recorded. For instance, compared to the White British ethnicity, subsequent records are more likely to be inconsistent for ethnic minorities, and are more often recorded as ‘other’, ‘not stated’, or ‘not known’ [40].

How ethnic minorities are categorised also affects the granularity with which health outcomes for different ethnic groups can be studied. Health-related data in the UK (e.g. [41]) often records ethnicity in accordance with the classification system recommended by ONS [42]. Any classification system inevitably focuses research on ethnic minorities for whom there is a category, while either neglecting ethnic minorities for whom there is no specific designation or grouping them together.

Grouping ethnic minorities together may be useful for identifying broad patterns arising from common experiences of discrimination and/or social exclusion. However, doing so may also obscure the extent of inequalities arising from within-group differences, which can be shaped by intersecting identities arising from different social, economic, and cultural contexts [43]. Lack of fine-grained data and small group sizes mean that the health and care needs of some ethnic groups are not well quantified. There is therefore an imperative to identify and optimise usage of data sources which do collect ethnic data in a meaningful way at a granular level.

A potential source of ethnic data is General Practice (GP) recording, which has improved in recent years. By 2024, over 95% of patients in England had a known ethnic category [44].

The North West London (NWL) Whole Systems Integrated Care (WSIC) contains valuable data from GP health records that can be used to quantify levels of health and use of care. The scale of the system means that even at smaller geographies (e.g. local authority level), minority groups can be studied at hitherto unprecedented scale. The aims of this paper are two-fold: (1) To describe the health status and use of healthcare and preventative services of the Romanian and Somali groups in Brent; and (2), to demonstrate that GP data can be useful for quantifying and describing the health conditions and utilisation of emerging communities.

## 2 Methods

### 2.1 Dataset

The NWL WSIC database contains linked, de-identified records from GPs, hospitals, and social care in NWL. The database also includes demographic information, such as birth month, sex, and index of multiple deprivation (IMD) decile - which is a small-area measure of relative deprivation across England and Wales. As of June 2025, there were records for 2,872,782 currently registered, living patients in the database.

Patients living in Brent, registered with a GP in the Brent health borough (previously Brent Clinical Commissioning Group, CCG), and alive during 1 April 2023 to 31 March 2024 formed our analytical sample.

### 2.2 Measures

### 2.3 Ethnicity

To categorise residents of Romanian and Somali ethnicity, GP records from 2010 onwards were searched for any entry containing the case-insensitive string “Romania” and “Somali” respectively. All other residents were categorised as “Rest of Brent”.

#### 2.3.1 Long-Term Conditions (LTCs)

The presence of LTCs was determined from GP records. We included 19 LTCs (Table 2) that are captured in the Quality Outcomes Framework or commonly included in descriptive studies of LTCs and recorded as (Systematized Nomenclature of Medicine – Clinical Terms) SNOMED CT codes.

#### 2.3.2 Healthcare Utilisation

Four measures were used as indicators for healthcare utilisation: number of GP encounters, number of emergency department attendances, number of avoidable emergency department attendances, and number of hospital admissions. Number of GP encounters was proxied by number of distinct days with an entry in the GP record. While this included administrative entries where there were no patient-GP interactions, this over-counting should nonetheless be similar across all ethnic groups. Similarly, number of emergency department attendances and number of hospital admissions were proxied by number of start dates. Emergency department attendances with a Healthcare Resource Group (HRG) code (VB11Z) indicating no further investigation or significant medication were labelled as an “avoidable” attendance – in that the issue could potentially have resolved in primary care [45]. These measures were calculated for the period from 1 April 2023 to 31 March 2024.

#### 2.3.3 Preventative Healthcare

Immunisation uptake and NHS health checks were flagged from SNOMED CT codes in GP records.

However, some childhood immunisations require multiple doses (e.g. MMR). Records do not always distinguish which dose was given. Since a minimum four-week interval is recommended between successive doses of the same vaccine [46], unless dose number was stated – or if doses were recorded as ‘booster’ doses – only records for the same individual made four weeks or more apart were regarded as separate, consecutive doses.

### 2.4 Statistical Analyses

To compare the prevalence of long-term conditions in Romanians and Somalis to the rest of Brent, age- and sex-standardised ratios were calculated. Age- and sex-specific rates of the rest of Brent were applied to the Romanian and Somali groups to derive expected counts, which were then compared against observed counts to derive the ratios.

A log-transformation (plus one) was applied to number of GP encounters, because the data demonstrated a positive skew. A linear regression model was then built to model the association between the transformed measure and ethnicity (Romanian, Somali, rest of Brent), adjusting for age, sex, deprivation in the area of residence, and number of LTCs.

For number of emergency department attendances, avoidable emergency department attendances, and hospital admissions, the measures were each binarised, creating two new measures indicating whether each patient had had one attendance (admission) or not, and whether they had multiple attendances (admissions) or not. A separate logistic regression was created to model the association between each of these six binary measures with ethnicity (Romanian, Somali, rest of Brent), also adjusting for age, sex, deprivation in the area of residence, and number of LTCs. Additionally, for the avoidable emergency department attendances regression models, patients who had not had any emergency department attendances were excluded from the model sample.

## 3 Results

### 3.1 Demographics

The search yielded 387,149 residents in Brent, out of whom 14,562 were categorised as Romanian and 13,446 were categorised as Somali. In the 2021 Census, 12,610 residents identified as Somali or Somalilander, while 17,722 residents identified as being Romanian-born [47]. It is worth mentioning that discrepancies may arise where residents are unregistered, or registered with GPs outside of Brent.

The proportion of males was highest in the Romanian group (54%), followed by the rest of Brent (52%), with the Somali group having the lowest proportion of males (48%). While median age was lowest for the Somali group (*Mdn*_Somali_ = 29, *Mdn*_Romanian_ = 34, *Mdn*_Other_ = 36), the interquartile range for the Romanian group was the smallest, and their age distribution was bimodal (see Appendix A). A greater proportion of the Somali group live in more deprived localities – with 52% of the group living in IMD deciles 1 and 2, compared to 14.7% of the Romanian group, and 18.9% of the rest of Brent (Table 1).

**Table 1.** Demographics

| Characteristic | Rest of Brent | Romanian | Somali |
| --- | --- | --- | --- |
| **N** | 359,141 | 14,562 | 13,446 |
| **Sex** |  |  |  |
| Female | 171,697 (48%) | 6,701 (46%) | 6,934 (52%) |
| Male | 187,444 (52%) | 7,861 (54%) | 6,512 (48%) |
| **IMD Decile<sup>1</sup>** |  |  |  |
| 1 - Most Deprived | 21,219 (5.9%) | 394 (2.7%) | 2,865 (21%) |
| 2 | 46,950 (13%) | 1,680 (12%) | 4,150 (31%) |
| 3 | 66,584 (19%) | 2,579 (18%) | 2,621 (19%) |
| 4 | 94,506 (26%) | 2,357 (16%) | 2,264 (17%) |
| 5 | 72,835 (20%) | 3,387 (23%) | 1,016 (7.6%) |
| 6 | 35,065 (9.8%) | 2,409 (17%) | 317 (2.4%) |
| 7 | 12,921 (3.6%) | 1,150 (7.9%) | 139 (1.0%) |
| 8 | 9,061 (2.5%) | 606 (4.2%) | 74 (0.6%) |

<sup>1</sup>Decile 10 represents the least deprived areas in England and Wales, but none of the localities in Brent fall within Deciles 9 or 10.

**Table 2.**
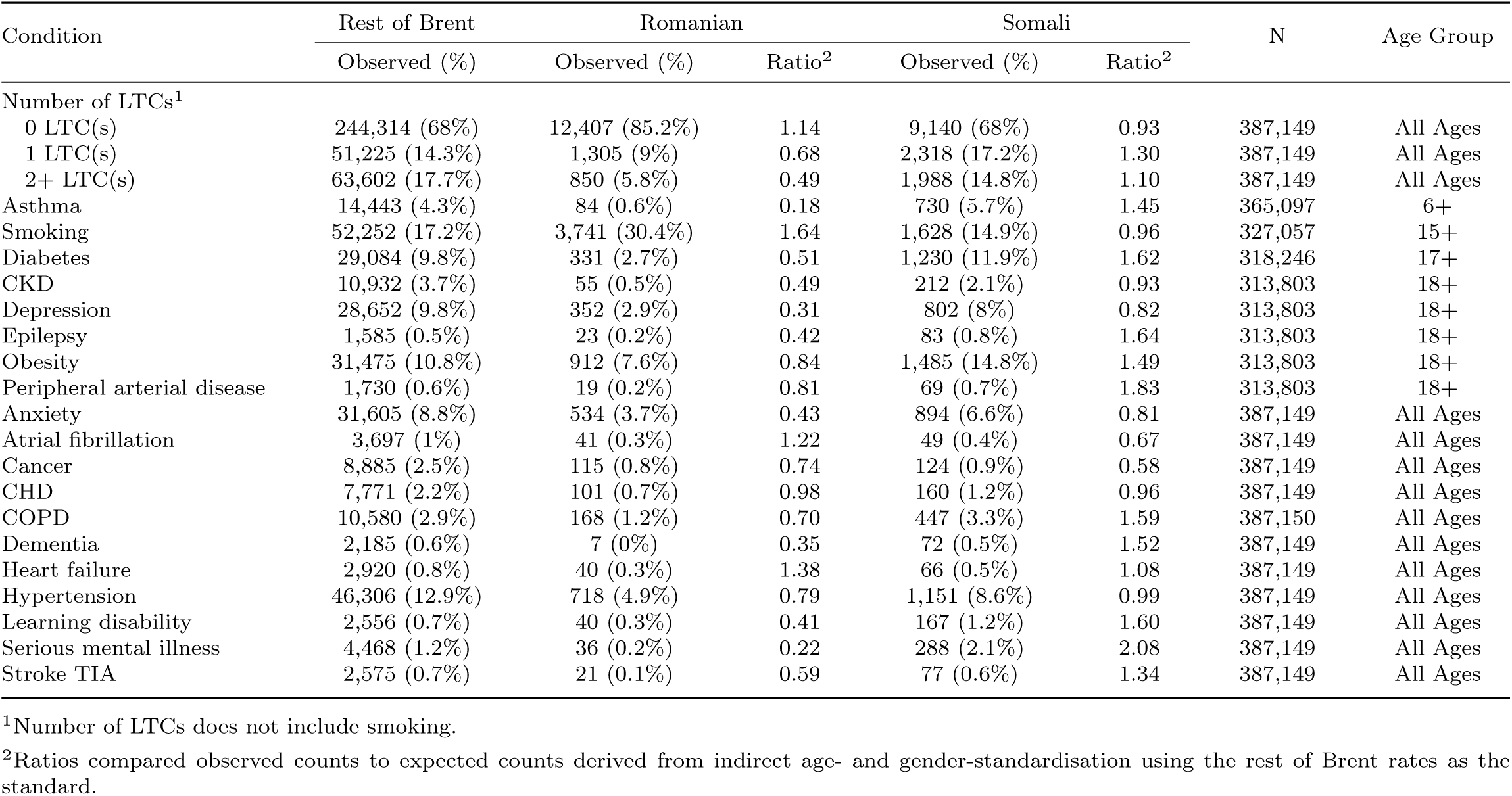
Health status by ethnicity

After accounting for age and sex differences in population structure, when compared to the rest of Brent, Somalis had higher prevalence of LTCs, and multiple LTCs, while Romanians had lower prevalence for both.

Despite a 30% prevalence of smoking, Romanians had lower prevalence of asthma, and COPD, compared to the rest of Brent. Indeed, other than smoking, the Romanian group only had higher prevalences for atrial fibrillation and heart failure.

Compared to the rest of Brent, Somalis have a higher prevalence of asthma and COPD, despite a slightly lower prevalence of smoking. Prevalence of obesity is higher as well, along with related diseases including diabetes, peripheral arterial disease, heart failure, and stroke. While prevalence of depression and anxiety are lower, the number with severe mental illness was more than double the age- and sex-standardised expected count.

### 3.2 Healthcare Usage

#### 3.2.1 Adults

In unadjusted analyses, Romanian adults generally had lower healthcare utilisation, with fewer GP encounters, and a lower proportion with emergency department attendances, hospital admissions, or flu vaccinations, compared to their rest of Brent counterparts of the same sex. For NHS health checks, completion rates were below 20% for all groups, and lowest for Romanian males (5.2%). A larger proportion of Romanian adults with an emergency department attendance had an avoidable emergency department attendance, compared to other adults (Table 3).

**Table 3.**
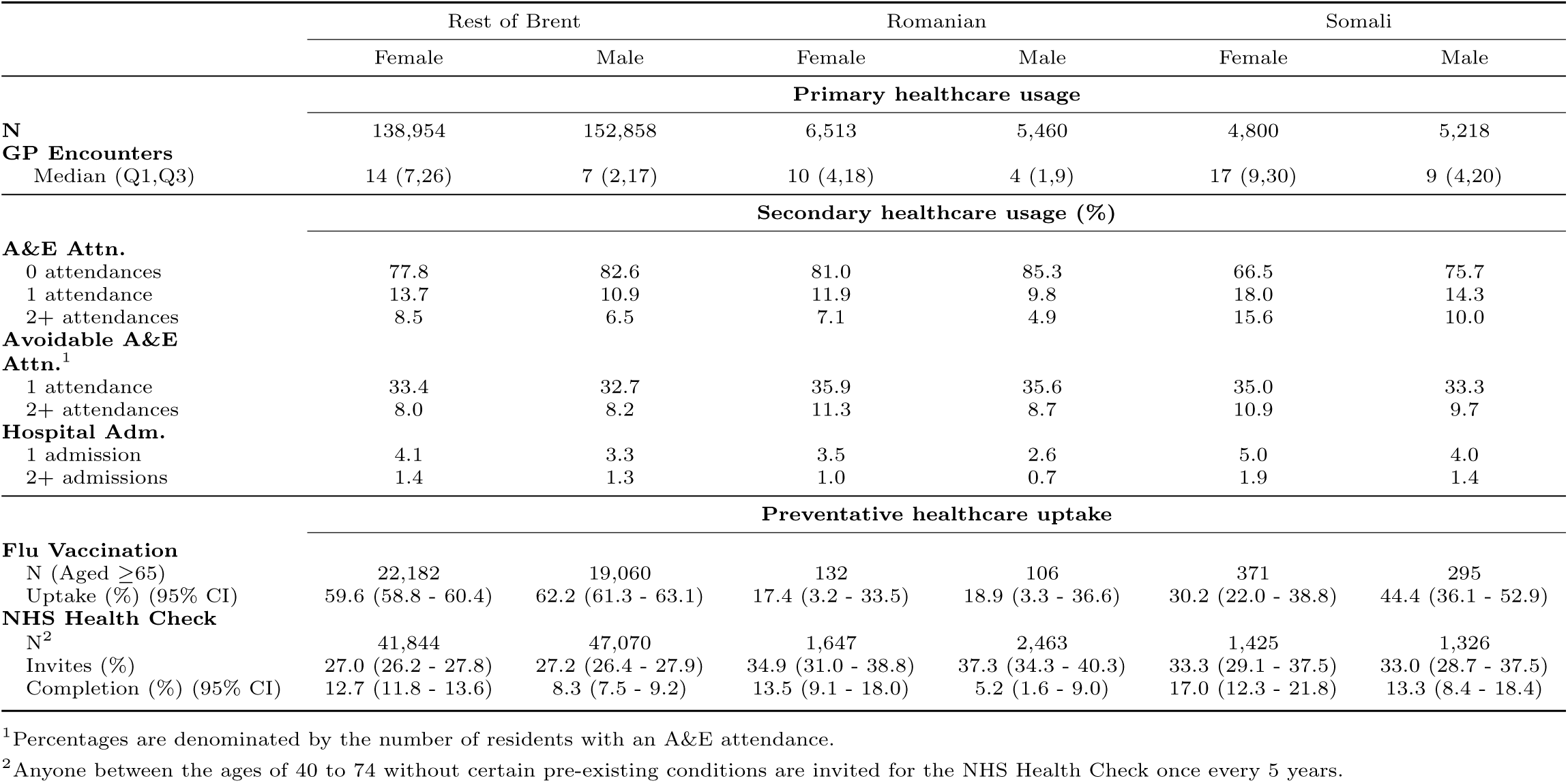
Healthcare usage for adults by ethnicity and sex

In contrast, Somali adults had higher healthcare utilisation across all measures than their rest of Brent counterparts of the same sex, with the exception of for flu vaccinations (Table 3). In particular, 15.6% - or almost 1 in 6 - of Somali women had multiple emergency department attendances, compared to 8.5% of rest of Brent women or 10.0% of Somali men.

After adjusting for age, sex, deprivation, and presence of LTCs, Romanians had significantly fewer GP encounters (*β* = -0.04; 95% CI: -0.04, -0.03; *t* = -10.87; *p < .*001) compared to the rest of Brent. Romanians were more likely to have had an avoidable emergency department attendance (OR: 1.10, 95% CI: 1.00 - 1.20), multiple avoidable emergency department attendances (OR: 1.32, 95% CI: 1.13 - 1.53), a hospital admission (OR: 1.30, 95% CI: 1.17 - 1.43), and multiple hospital admissions (OR: 1.30, 95% CI: 1.05 - 1.59) (Table 4).

**Table 4.**
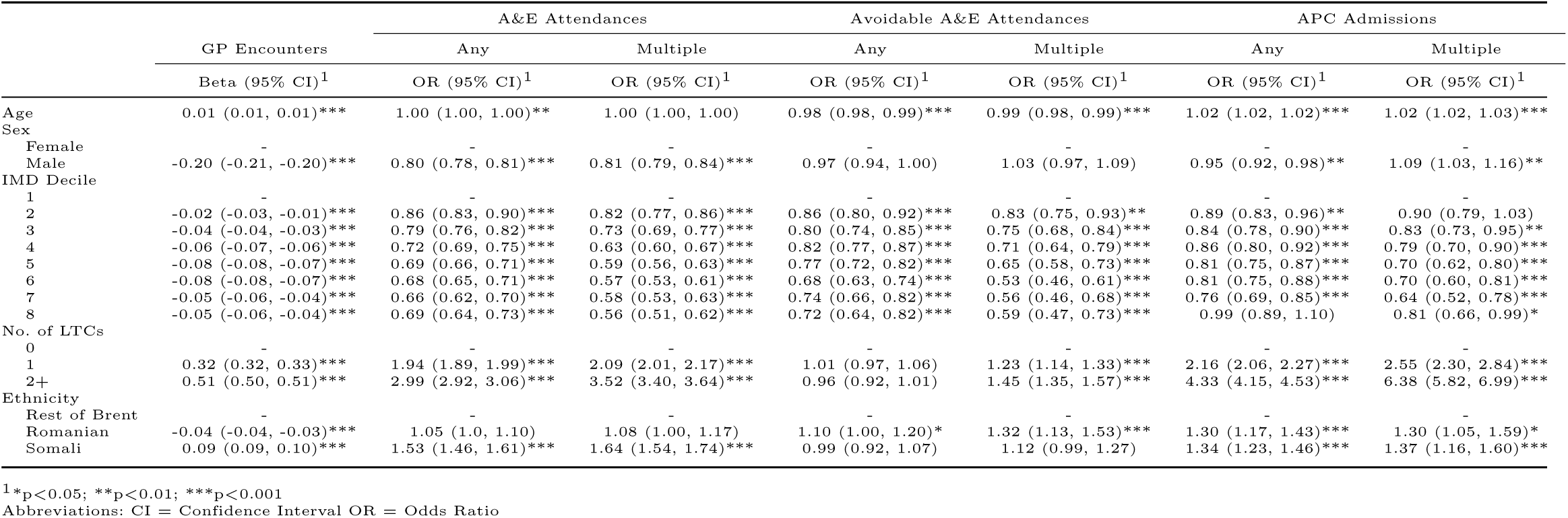
Association between healthcare utilisation and ethnicity among adults in Brent.

In contrast, Somalis had significantly more GP encounters (*β* = -0.04; 95% CI: 0.09, 0.10; *t* = 24.29, *p < .*001). Somalis were also more likely to have an emergency department attendance (OR: 1.53, 95% CI: 1.46 - 1.61), multiple emergency department attendances (OR: 1.64, 95% CI: 1.54 - 1.74), a hospital admission (OR: 1.34, 95% CI: 1.23 - 1.46), and multiple hospital admissions (OR: 1.37, 95% CI: 1.16 - 1.60) (Table 4).

#### 3.2.2 Children (Under 18s)

In unadjusted analyses, Romanian children had fewer GP visits but a larger proportion of Romanian children with an emergency department attendance also had an avoidable emergency department attendance. Immunisation uptake for Romanian children was substantially lower across all milestones compared to the rest of Brent (Table 5).

**Table 5.** Healthcare usage for children (under 18 years) by ethnicity and sex

|  | Rest of Brent |  | Romanian |  | Somali |  |
| --- | --- | --- | --- | --- | --- | --- |
|  | Female | Male | Female | Male | Female | Male |
| **Primary healthcare usage** |  |  |  |  |  |  |
| **N** | 32,743 | 34,586 | 1,241 | 1,348 | 1,716 | 1,712 |
| **GP Encounters** |  |  |  |  |  |  |
| Median (Q1,Q3) | 6 (2,11) | 6 (2,11) | 4 (1,10) | 5 (2,11) | 6 (2,11) | 6 (2,12) |
| **Secondary healthcare usage (%)** |  |  |  |  |  |  |
| **A&E Attn.** |  |  |  |  |  |  |
| 0 attendances | 77.0 | 73.7 | 75.8 | 75.1 | 73.8 | 71.2 |
| 1 attendance | 14.5 | 16.1 | 14.1 | 14.3 | 16.0 | 15.8 |
| 2+ attendances | 8.6 | 10.2 | 10.1 | 10.6 | 10.2 | 13.0 |
| **Avoidable A&E Attn.<sup>1</sup>** |  |  |  |  |  |  |
| 1 attendance | 44.0 | 44.4 | 45.3 | 50.0 | 41.2 | 38.5 |
| 2+ attendances | 15.2 | 15.0 | 21.7 | 21.4 | 14.0 | 16.8 |
| **Hospital Adm.** |  |  |  |  |  |  |
| 1 admission | 3.1 | 3.6 | 2.3 | 3.6 | 3.5 | 5.0 |
| 2+ admissions | 0.7 | 1.0 | 0.6 | 0.5 | 0.9 | 1.1 |
| **Immunisation uptake (%)<sup>2</sup>** |  |  |  |  |  |  |
| Rotavirus (12m) | 85.9 (84.7 - 87.0) |  | 59.4 (50.9 - 67.4) |  | 78.6 (68.7 - 86.0) |  |
| PCV Dose 1 (12m) | 91.6 (90.6 - 92.5) |  | 72.9 (64.8 - 79.8) |  | 88.1 (79.5 - 93.4) |  |
| PCV Booster (24m) | 81.3 (79.9 - 82.5) |  | 59.9 (52.2 - 67.1) |  | 57.5 (48.6 - 66.0) |  |
| 6-in-1 (12m) | 87.4 (86.2 - 88.5) |  | 63.9 (55.5 - 71.6) |  | 79.8 (70.0 - 87.0) |  |
| 6-in-1 (24m) | 85.3 (84.1 - 86.5) |  | 71.6 (64.2 - 78.0) |  | 80.0 (72.0 - 86.2) |  |
| MMR1 (24m) | 83.4 (82.1 - 84.6) |  | 63.6 (55.9 - 70.6) |  | 54.2 (45.3 - 62.8) |  |
| MMR1 (5y) | 87.4 (86.3 - 88.5) |  | 72.5 (65.6 - 78.5) |  | 65.1 (56.6 - 72.8) |  |
| MMR2 (5y) | 78.9 (77.5 - 80.2) |  | 50.0 (42.8 - 57.2) |  | 41.1 (33.0 - 49.7) |  |
| Hib/MenC booster (24m) | 82.3 (81.0 - 83.5) |  | 63.6 (55.9 - 70.6) |  | 60.8 (51.9 - 69.1) |  |
| Hib/MenC booster (5y) | 83.7 (82.4 - 84.9) |  | 68.1 (61.0 - 74.5) |  | 59.7 (51.1 - 67.8) |  |

<sup>1</sup>Percentages are denominated by the number of residents with an A&E attendance.
<sup>2</sup>Appendix B provides a comparison of immunisation rates derived from the WSIC database to official statistics.

For Somalis, a higher proportion of male children had had multiple emergency department attendances compared to their rest of Brent counterparts. However, of the children with an emergency department attendance, a smaller proportion of them had an avoidable emergency department attendance. Somali children also had lower immunisation uptake across all milestones. However, where Romanian children tended to have consistently poor uptake for all milestones, differences in uptake rates between Somali children and rest of Brent children were smaller for immunisations scheduled for the first 3 or 4 months after birth (i.e. Rotavirus, PCV Dose 1, DTap-IPV-Hib-HepB) and larger for immunisations taken outside this time period (i.e. PCV Booster, MMR1, MMR2, and Hib/MenC) (Table 5).

After adjusting for age, sex, deprivation, and presence of LTCs, Romanian children had significantly fewer GP encounters (*β* = -0.05; 95% CI: -0.06, -0.03; *t* = -6.19, *p < .*001), compared to their rest of Brent counterparts. Romanian children were also more likely to have had an avoidable emergency department attendance (OR: 1.48, 95% CI: 1.25 - 1.77), and multiple avoidable emergency department attendances (OR: 1.51, 95% CI: 1.26 - 1.87). These trends were similar to their adult counterparts. However, Romanian children were also more likely to have had multiple emergency department attendances (OR: 1.17, 95% CI: 1.02 - 1.33) (Table 6).

**Table 6.**
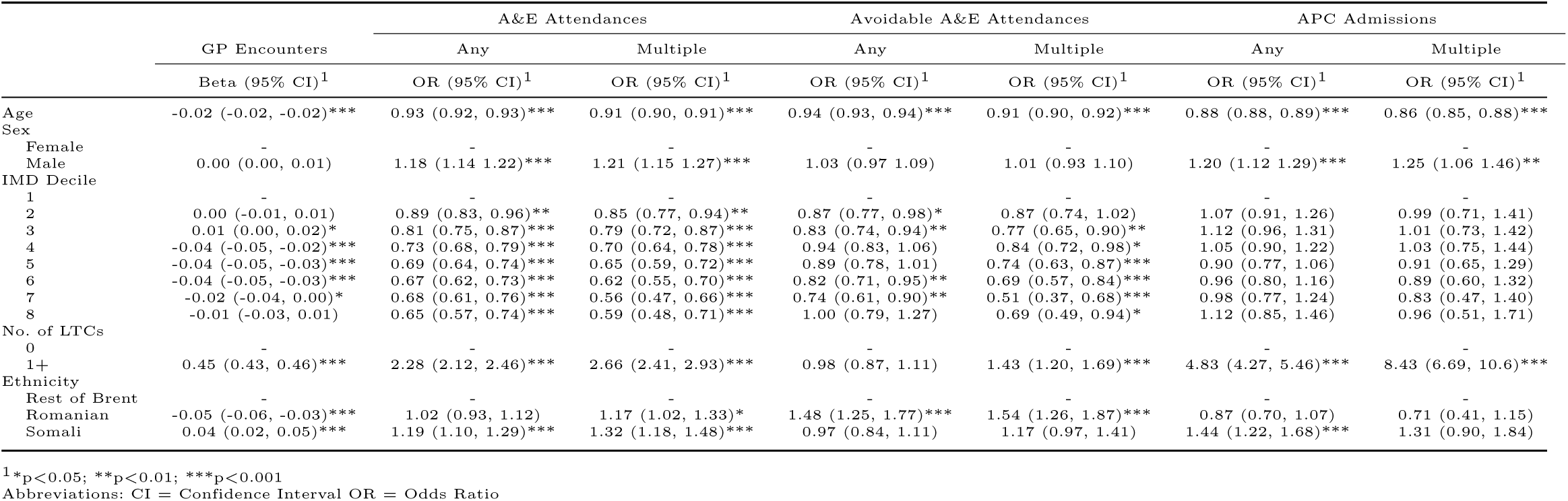
Association between healthcare utilisation and ethnicity among Under 18s in Brent.

In contrast, Somali children had significantly more GP encounters (*β* = 0.04; 95% CI: 0.02, 0.05; *t* = 5.37, *p < .*001). Like Somali adults, Somali children were more likely to have had an emergency department attendance (OR: 1.19, 95% CI: 1.10 - 1.29), multiple emergency department attendances (OR: 1.32, 95% CI: 1.18 - 1.48), and a hospital admission (OR: 1.44, 95% CI: 1.22 - 1.68) (Table 6).

## 4 Discussion

Based on diagnoses recorded in general practice, LTC prevalence is lower in the Romanian community compared with the rest of Brent, although smoking prevalence is higher. Controlling for age, sex, deprivation and LTCs, the use of secondary healthcare services is higher for Romanian adults, while use of emergency department services is higher for Romanian children.

LTC prevalence for the Somali community is similar to the rest of Brent, although they experience a higher prevalence of obesity and some other LTCs. Adjusted analysis showed higher use of both primary and secondary healthcare services compared with the rest of Brent.

### 4.1 LTC Prevalence

While Romanians have lower LTC prevalence of diagnosed LTCs, it is unclear whether this reflects true LTC prevalence or is a product of lower healthcare utilisation. One explanation for true lower LTC prevalence is the ‘Healthy Migrant’ effect. However, our findings may also reflect undiagnosed or unrecorded cases as a consequence of fewer encounters with local healthcare services and fewer opportunities for diagnosis. As evidence of the latter, [17] notes Romanian preference for returning to Romania for treatment. Additionally, while disease prevalence is generally lower for Romanians than the rest of Brent - even for asthma and COPD despite higher rates of smoking - prevalence of diseases with urgent and acute presentations (i.e. atrial fibrillation and heart failure) are higher, and may be more likely to reflect actual prevalence.

Comparatively, there is no previous literature describing substantial numbers of people from Somali communities travelling outside the UK for healthcare and so recorded disease prevalence is likelier to reflect actual prevalence. The exception to this, however, may be mental illness. Despite lower prevalence of depression and anxiety, the rate of serious mental illness is more than double the age- and sex-standardised expected rate. Given reports of stigmatisation and low awareness of mental health and disease amongst Somalis [34, 36, 37], high prevalence of serious mental illness may be partly due to under-diagnosis and under-treatment earlier in the disease pathway.

Prevalence of obesity is higher in the Somali community. Previous studies suggest that healthy body weight [25, 26] - and health awareness, such as what constitutes a healthy diet [23, 26] or physical activity [24] could contribute to this. Together, these are likely to also be related to the relatively higher prevalences of diabetes, peripheral arterial disease, heart failure, and stroke.

### 4.2 Healthcare Usage

Romanian adults had fewer GP encounters and higher odds of secondary healthcare usage (one or more avoidable emergency department attendances, having at least one admission, and having multiple admissions). This corroborates reports by healthcare providers that Romanian community members are more likely to access acute emergency care than primary care, possibly because of differences in the healthcare system organisation, entitlements, or the emphasis on prevention and early intervention [20]. We may, however, have underestimated these differences because our analyses do not include Romanians who are unregistered at GPs. This is important to note because higher emergency department attendance tends to be associated with unregistered residents [18, 48].

While Romanians overall had fewer diagnosed LTCs, compared to rest of Brent counterparts with the same number of LTCs, Romanians were more likely to have had a hospital admission. Whether admissions were elective or otherwise, and reasons for admission were not specified in this study. However, this could possibly be another manifestation of undiagnosed or unrecorded LTCs, as a result of fewer GP visits by Romanians.

Similarly, Romanian children had fewer GP encounters, and higher odds of secondary healthcare utilisation (multiple emergency department attendances, and one or more avoidable emergency department attendances). The difference in odds of avoidable emergency department attendance between Romanian children and the rest of Brent is larger than the difference between the respective adult counterparts. Under-estimation due to non-registration at the GPs may be less common in children. A 2022 report on the duration of migrant stays within the UK found that work migrants who bring children with them are more likely to stay in the UK long term [49]. Taken together, Romanians with children may be more likely than their childless counter-parts to decide to stay within the UK, and engage with local healthcare - starting with GP registration. If so, a greater proportion of Romanian children (compared to adults) would be registered, allowing their hospital records to be linked, retrieved, and reflected in the results of the analysis here.

For Somalis, even after adjusting for the presence of long-term conditions and deprivation, both children and adults have higher healthcare utilisation across most measures. Avoidable emergency department attendance is the exception to this, suggesting that health needs may more often drive emergency healthcare seeking in this group.

#### 4.2.1 Child Immunisation Uptake

Vaccination uptake for MMR is particularly low in Somali children compared to the rest of Brent. This corroborates reports of Somali suspicion toward MMR vaccination [28, 29] and is similar to reported MMR1 uptake among Somalis in Tower Hamlets (56%) [30]. Considering that the latter analysed data from 2009 to 2011, the similarity in MMR vaccination uptake by Somalis from that time period to this implies a distressing lack of progress in dispelling the autism myth.

While uptake of the DTaP-IPV-Hib-HepB vaccine is higher than for MMR in Somali children (79.8% at one year of age, 80.0% at two years of age), these rates are lower than reported in literature (∼85% at one year of age [31, 32], ∼88% at two years of age [32]). However, the lower rate for Somali children in Brent may reflect the fact that overall uptake rate of the diphtheria-containing vaccines is lower in Brent compared to the cohorts in the reported literature.

Although uptake rates are lower for Somali children compared to the rest of Brent across all milestones, the degree of disparity in uptake rates appears to vary depending on the timing of vaccine administration. The difference between Somali children and rest of Brent children in uptake rates of vaccinations administered within months of birth (i.e. DTaP-IPV-Hib-HepB, Rotavirus, PCV Dose 1) was smaller than the difference for vaccinations administered 1 year or more after birth (i.e. MMR, Hib/MenC, PCV Booster). This suggests that at least up to 4 months, Somali children are relatively well-tracked and followed-up within the NHS system. Thus, the challenge - and where efforts should be targeted at - is consistently following up with Somali parents 1 year and more after birth.

Previous studies on possible reasons for lower vaccination uptake in Romanian children have pointed to language barriers that hinder both Romanians from obtaining credible medical information [17, 20] and healthcare providers from obtaining complete and accurate vaccination histories [17]. Differences in the vaccination schedules of Romania and the UK, as well as the types of vaccinations available (or not) was a source of confusion as well [17]. The system of appointment booking was thought to impede uptake - healthcare providers suggested that drop-in sessions could potentially be more effective [17]. For example, a vaccination event held in tandem with a community meal at an established, trusted, community centre with translated medical information was lauded as an example of a successful approach toward attaining better immunisation rates [21].

#### 4.2.2 Adult Preventative Health

Flu vaccination uptake was lowest for Romanians. Previous studies found confidence in natural immunity was a primary motivator in low vaccine uptake in the general population [50], while healthcare workers themselves were also less positive regarding vaccine benefits compared to other European counterparts [51].

For NHS Health Checks, Somalis had higher completion rates, while Romanians had lower completion rates, compared to the rest of Brent. These differences may be related to healthcare utilisation in general, especially since NHS Health Checks are delivered by GPs. Additionally, while the NHS Health Check does not include people with various chronic conditions^1^, the high prevalence of chronic conditions in the community may nonetheless motivate Somalis with no pre-existing conditions to complete the checks.

^1^The rationale behind the exclusion criteria is that those with these chronic conditions should be receiving regular check-ups already. A full list of the exclusion criteria can be found at https://www.nhs.uk/ conditions/nhs-health-check/

### 4.3 Limitations

Data quality is contingent on the correctness and completeness of GP recording. There is considerable overlap in the GPs all ethnic groups in our study are registered at, which should minimise differences, although differences in how GPs identify and record symptoms and conditions for different ethnic groups cannot be ruled out.

Our analysis does not capture healthcare data from unregistered people, who arguably shoulder the greatest health burden. Nonetheless, it is sufficient to demonstrate that people from minority ethnic groups who do register with GPs experience disproportionately poorer health status. While it must be acknowledged that unregistered and registered people may not necessarily be a homogenous group, it is hoped that some of the findings in this paper can be used to drive improvements that would reduce health burdens of these emerging communities as a whole.

The extent of under-registration, however, likely varies between communities. For example, qualitative evidence suggests that some Romanian migrants return to Romania for healthcare [16–18], which may reduce both their engagement with preventative services in the UK and their likelihood of GP registration. Furthermore, while the number of registered patients in the rest of Brent and among the Somali population exceeded corresponding 2021 Census figures, the number of registered Romanian patients was lower. This discrepancy strengthens the likelihood that under-registration disproportionately affects the Romanian population, and suggests that the analysis may underestimate the extent of need in this group.

## 5 Conclusion

In summary, Somalis experience poorer health status, and have higher healthcare use, even after health conditions are accounted for. In contrast, Romanians appear to be in better health, and have lower primary healthcare use. However, the latter may be a product of undiagnosed or unrecorded LTCs, and when accounting for the number of LTCs, Romanian adults instead have higher odds of having had hospital attendance(s). While both Romanian and Somali children have lower immunisation uptake, areas for improvement may differ. For instance, for Somalis, uptake of immunisations within the first few months of life are higher - suggesting efforts should be focused on following up. This is especially the case since it is noted that Somalis do engage with other preventative services, such as the NHS Health Checks. Comparatively, the area of focus for Romanians may be supporting engagement with primary healthcare. Unlike Somali children, uptake for Romanian children is lower across all immunisations - not just those administered a year or more after birth. Addressing language barriers may be key to efforts in both the Somali [33] and Romanian [17, 18, 20, 21] communities. Even within a relatively small geography, this paper demonstrates that ethnic minority health care and outcomes can be quantified with reasonable scale using GP data. This can be extended to other emerging communities, who do not have a designated category in typical ethnic classification, but have corresponding SNOMED CT codes (e.g. Brazilians).

Finally, this analysis has demonstrated that GP health records can be leveraged for granular monitoring. These are a valuable resource - especially at smaller geographies - to provide evidence and insight for efforts to tackle health inequalities experienced by emerging communities closer to the ground.

## Declarations

### 6.1 Ethics approval and consent to participate

Ethics approval was not required for this secondary analysis of de-identified patient data. Approval for this study was provided by the WSIC Care Data Access Committee.

### 6.2 Consent for publication

Not applicable.

### 6.3 Data availability

Access to data to replicate the study can be requested from the WSIC Data Access Committee. https://www.nwlondonicb.nhs.uk/professionals/ whole-systems-integrated-care-wsic/information-de-identified-users

### 6.6 Authors’ contributions

John Licorish (JL) and Mai Stafford (MS) conceptualised the research idea. All authors contributed to designing and interpreting the study. Sarah Yeoh (SY) conducted the data analysis, with support from MS. SY produced the draft of the paper, and MS critically revised the manuscript. All authors read and approved the final manuscript.

## Acknowledgments

We thank the North West London Whole Systems Integrated Care team for their continued support with metadata and data access.

This work uses data provided by patients and collected by the NHS as part of their care and support.

## Appendix A Age Distribution by Sex and Ethnicity

**Fig. A1.**
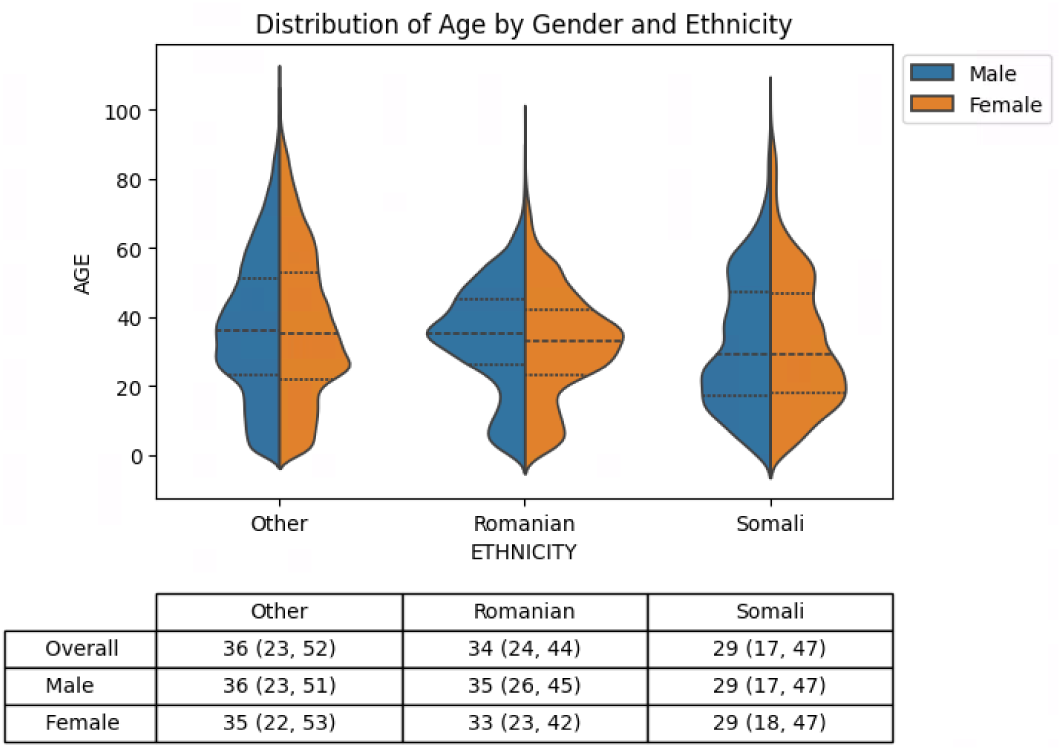
Age distribution by sex and ethnicity. Medians (Quartile 1, Quartile 3) are presented in the table.

## Appendix B Obtained Rates and Official Estimates

For all milestones less MMR1 at 5 years, the uptake rates obtained from the WSIC database fall within the confidence intervals of NHS England rates, demonstrating the reliability of figures derived from the database (Table B1).

**Table B1.** A comparison of Brent population immunisation rates derived from WSIC data and from NHS England statistics.

|  | NHS England (95% CI) | WSIC |
| --- | --- | --- |
| Rotavirus (12mo) | 85.2% (84.1, 86.2) | 84.7% |
| PCV Dose 1 (12mo) | 90.5% (89.6, 91.4) | 90.8% |
| PCV Booster (24mo) | 79.3% (78.0, 80.5) | 79.5% |
| DTaP-IPV-Hib-HepB (12mo) | 86.3% (85.3, 87.3) | 86.4% |
| DTaP-IPV-Hib-HepB (24mo) | 85.4% (84.3, 86.4) | 84.6% |
| MMR1 (24mo) | 81.6% (80.4, 82.7) | 81.7% |
| MMR1 (5y) | 84.8% (83.7, 85.8) | 86.0% |
| MMR2 (5y) | 75.0% (73.7, 76.3) | 76.3% |
| Hib/MenC booster (24mo) | 81.2% (80.0, 82.3) | 80.8% |
| Hib/MenC booster (5y) | 82.4% (.) | 82.1% |

## 6 List of abbreviations

A&E: Accident and emergency
APC: Admitted patient care
CCG: Clinical Commissioning Group
COPD: Chronic obstructive pulmonary disease
DTaP-IPV-Hib-HepB: Diphtheria, tetanus, pertussis, inactivated Poliovirus vaccine, Haemophilus influenzae type b, and hepatitis B
GP: General practice
Hib/MenC: Haemophilus influenzae type b/meningococcal group C
HRG: Healthcare Resource Group
IMD: Index of Multiple Deprivation
LTC: Long-term condition
MMR: Measles, mumps and rubella
NHS: National Health Service
NWL: North West London
ONS: Office for National Statistics
PCV: Pneumococcal conjugate vaccine
SNOMED CT: Systematized Nomenclature of Medicine – Clinical Terms
UK: United Kingdom
WSIC: Whole Systems Integrated Care

